# Laboratory findings that predict a poor prognosis in COVID-19 patients with diabetes: A meta-analysis

**DOI:** 10.1101/2020.07.02.20145391

**Authors:** Francisco Alejandro Lagunas-Rangel, Venice Chávez-Valencia

## Abstract

Diabetes is one of the main comorbidities in patients infected with the SARS-CoV-2 virus, the causative agent of the new coronavirus disease 2019 (COVID-19). Because the presence of diabetes and COVID-19 in the same patient is related to a poor clinical prognosis and a high probability of death, it is necessary to determine what findings allow us to predict a good or bad resolution of the disease in order to opt for a traditional treatment or a more incisive one. In this way, in the present work we analyze which laboratory parameters showed differences in patients with COVID-19 and diabetes who recovered and in those who had complications or died.

## Introduction

The new coronavirus disease 2019 (COVID-19) is caused by an infection with the SARS-CoV-2 coronavirus. Severe cases of COVID-19 can rapidly progress to acute respiratory distress syndrome (ARDS), septic shock, multiple organ dysfunction syndrome (MODS), and ultimately patient death.^1^ In COVID-19 patients, diabetes mellitus (DM) is one of the main comorbidities associated with severe disease, ARDS, and increased mortality.^2^ This relationship is due, in part, to the fact that patients with diabetes have a permanent pro-inflammatory state that makes them more susceptible to an inflammatory cytokine storm, often their innate immune system is compromised, and they also have an underlying hypercoagulable pro-thrombotic state. Meanwhile, COVID-19 can worsen insulin resistance, making it difficult to control glucose in people with diabetes, and also promotes strong inflammatory and coagulation processes.^3,4^ Since establishing an early prognosis in patients with diabetes infected with SARS-CoV-2 could help in their treatment decisions, in the present work we analyzed which laboratory parameters showed differences that could predict a good or bad resolution of the disease.

## Material and Methods

We carry out an electronic search using different search engines such as Medline (PubMed interface), Scopus, Web of Science via Raven and Google Scholar, using the keywords “Diabetes + COVID-19” OR “Diabetes + SARS-CoV-2”, without any date (until June 30, 2020) or language restrictions. The titles, abstracts and full texts of all the articles identified according to these criteria were analyzed, considering for our meta-analysis only those that reported prognostic data in patients with COVID-19 and diabetes (defined as with and without the need to enter the intensive care unit, survivors and non-survivors or alive and deceased).

Only four studies were considered in our meta-analysis^5–8^ and only the laboratory parameters that appear in all articles were analyzed. The mean and standard deviation were extrapolated from the median, range, and sample size as previously described.^9^ We analyzed whether there are significant differences (p < 0.05) in the values of age, glycated hemoglobin (HbA1c), white blood cell (WBC) count, neutrophils (NEU), lymphocytes (LYM), platelets (PLT), D-dimer, cardiac troponin I (cTnI), alanine aminotransferase (ALT), creatinine (Cr), C-reactive protein (CRP), neutrophil-to-lymphocyte ratio (NLR), and procalcitonin (PCT) between patients who survived and those who presented complications or died. Meta-analysis was performed using Comprehensive Meta-Analysis Software version 3 (2013, Biostat, Englewood, NJ) calculating the standardized mean difference (SMD) and the 95% confidence interval (95% CI) of all parameters.

## Results and Discussion

All the studies were conducted in the Chinese population, considering a total of 303 patients with COVID-19 and diabetes, of whom 192 (63.36%) recovered without complications and 111 (36.63%) had complications or died. Samples between studies ranged from 28 to 153 patients. The SMD for each parameter is summarized in Figure 1. For most of the parameters, a random effects model was used since its heterogeneity (I^2^ statistics) exceeded 50%, with the exception of WBC and Cr where it was less and, therefore, a fixed effects model was used. The results of our analysis showed that HbA1c (SMD = 0.545, 95% CI = −1.001 to 2.091), PLT (SMD = −1.117, 95% CI = −2.553 to 0.318) and ALT (SMD = 0.752, 95% CI = −0.491 to 1.995) did not have significant differences between the two groups. Age (SMD = 1.686, 95% CI = 0.409 to 2.963), WBC (SMD = 2.002, 95% CI = 1.696 to 2.308), NEU (SMD = 2.472, 95% CI = 1.833 to 3.110), D-dimer (SMD = 1.930, 95% CI = 1.262 to 2.599), Cr (SMD = 1.482, 95% CI = 1.196 to 1.767), CRP (SMD = 3.244, 95% CI = 2.432 to 4.056), NLR (SMD = 3.165, 95% CI = 2.216 to 4.115), PCT (SMD = 1.402, 95% CI = 0.516 to 2.289), and cTnI (SMD = 1.342, 95% CI = 0.192 to 2.492) showed higher values in patients who had complications or died, while LYM (SMD = −2.577, 95% CI = −3.178 to −1.975) values were higher in patients who survived. Notably, CRP, NLR, NEU, LYM and WBC were the ones that presented the greatest differences between the groups.

**Figure 1.**
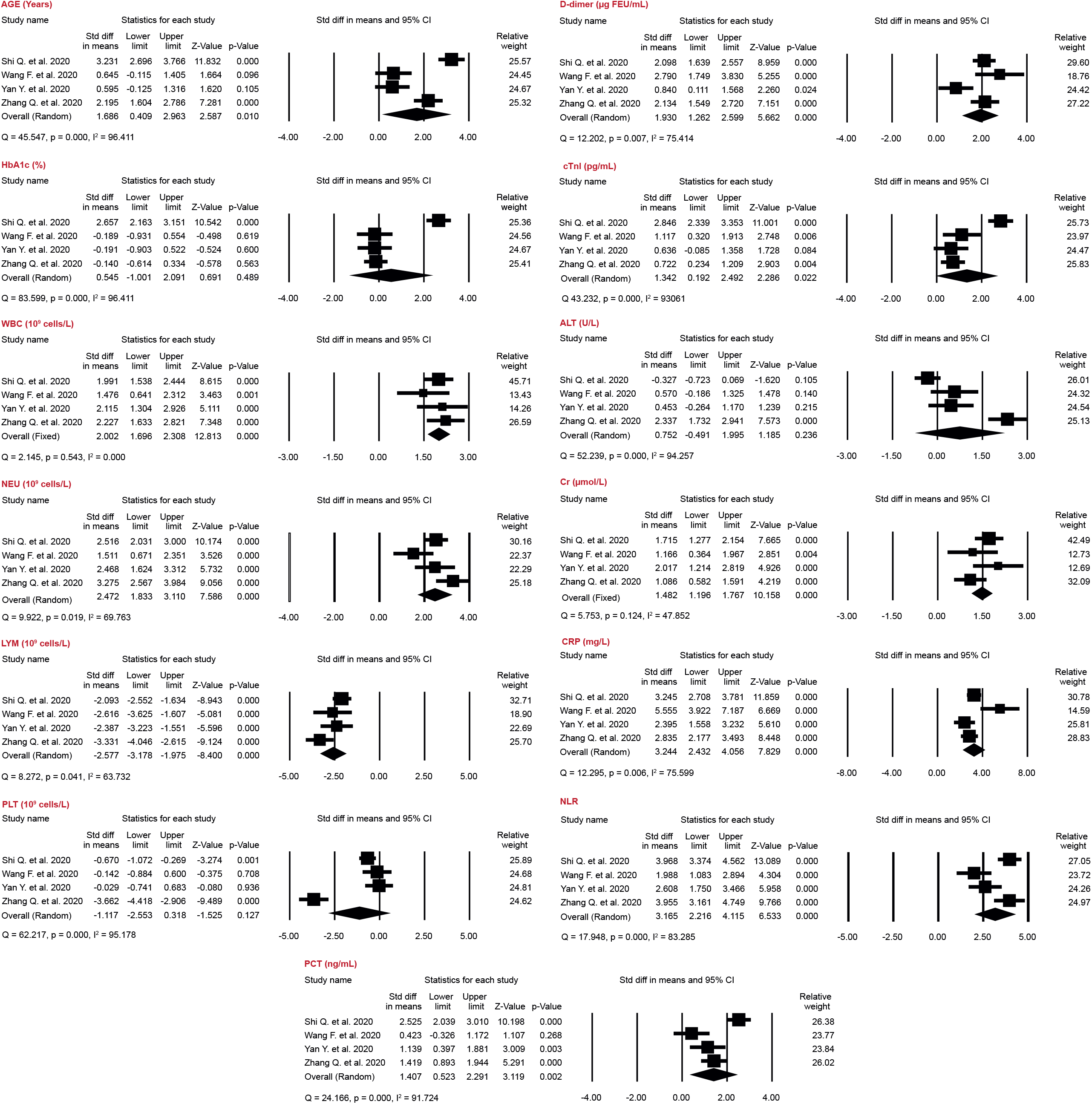
Forest plot of standardized mean difference (SMD) and 95% confidence interval (95% CI) of all the parameters analyzed. The units used in the analysis were: age (years), HbA1c (%), WBC (10^9^ cells/L), NEU (10^9^ cells/L), LYM (10^9^ cells/L), PLT (10^9^ cells/L), D-dimer (μg FEU/mL), cTnl (pg/mL), ALT (U/L), Cr (μmol/L), CRP (mg/L), and PCT (ng/mL).

Diabetes mellitus has been significantly associated with the mortality risk of patients with COVID-19.^10,11^ In general, patients with COVID-19 and diabetes who presented a poor prognosis showed higher levels of the inflammatory parameters, but at the same time an immunocompromise due to the significant reduction in the number of lymphocytes. Likewise, the parameters associated with coagulation and cardiac activity were increased in patients who had complications or died. No parameter could be identified that was clearly different between COVID-19 patients with diabetes and those without. Although blood glucose control has been reported to reduce the risk of death and detrimental complications^1^, we found no significant difference in HbA1c values, possibly due to good patient control prior to SARS-CoV-2 infection. The CORONADO study reported that prior glucose control does not have an impact on the severity of COVID-19 in people with diabetes.^12^ It is important to mention that the presence of obesity was not reported in any of the studies analyzed and the influence of BMI could not be investigated or included. Data on antidiabetic therapy at baseline and during treatment are not considered, and it is unknown what percentage of the population has uncontrolled diabetes.

## Data Availability

The analyzed data can be found in the referenced articles.

## Acknowledgements

FALR is recipient of a doctoral scholarship (Application number 2018-000012-01NACF- 07226) from the National Council of Science and Technology, CONACyT.

